# Quantitative assessment of collagen architecture from routine histopathological images shows concordance with Second Harmonic Generation microscopy

**DOI:** 10.64898/2026.03.31.26349841

**Authors:** Vedant Ingawale, Krishnendu Dandapat, Jyothsna Konkada Manattayil, Shashank Gupta, L S Shashidhara, Chaitanyanand Koppiker, Nameeta Shah, Varun Raghunathan, Madhura Kulkarni

## Abstract

Collagen organisation within the tumour microenvironment plays a critical role in tumour progression and has emerged as an important structural biomarker in cancer. Second Harmonic Generation (SHG) microscopy enables label-free visualisation and quantitative assessment of fibrillar collagen architecture; however, its high cost, specialised instrumentation, and limited field-of-view restrict routine clinical application. In this study, we evaluated whether collagen features quantified from digitally scanned Masson-Goldner’s Trichrome-stained histopathological sections can approximate measurements obtained from SHG microscopy.

Formalin-fixed paraffin-embedded breast tumour tissues, including benign and invasive ductal carcinoma (IDC) samples with varying collagen content, were analysed using SHG microscopy and whole-slide brightfield imaging. Matched regions of interest were analysed using two independent digital image analysis approaches: a conventional ImageJ-based workflow (TWOMBLI) and a machine learning-based computational pipeline. Collagen structural parameters including collagen deposition area, fibre number, and alignment metrics were quantified and compared across imaging modalities using correlation analysis.

SHG signals were consistently detected from trichrome-stained sections, confirming compatibility of SHG imaging. Quantitative comparison demonstrated significant concordance between SHG-derived collagen metrics and those obtained from digital image analysis pipelines, particularly for collagen area and fibre alignment.

These findings demonstrate that computational analysis of routine histopathological images can capture key spatial features of collagen organisation comparable to SHG microscopy. Digital pathology-based collagen quantification therefore, represents a scalable and clinically accessible approach for assessing extracellular matrix architecture in tumour tissues.

**Author Summary:** Quantitative assessment of extracellular matrix architecture remains a critical yet underutilized dimension of cancer pathology. While Second Harmonic Generation (SHG) microscopy provides a gold-standard, label-free approach to interrogate fibrillar collagen organisation, its limited accessibility has prevented integration into routine clinical workflows. This creates a translational gap between advanced imaging capabilities and scalable clinical implementation.

Here, we address this gap by demonstrating that collagen structural features derived from **routine histopathological whole-slide images** can approximate SHG-derived measurements. Using matched regions from breast tumour tissues, we perform a systematic cross-modality comparison between SHG microscopy and two independent computational pipelines, a conventional ImageJ-based workflow and a machine learning-based framework. We observe strong concordance across key spatial metrics, including collagen deposition, fibre density, and alignment.

Importantly, we establish that SHG imaging is fully compatible with trichrome-stained sections, enabling direct validation on identical tissue regions. Our findings show that widely available digital pathology data, when coupled with computational analysis, can recover biologically and clinically relevant features of collagen architecture previously accessible primarily through specialized optical systems.

This work introduces a **scalable, cost-effective, and clinically translatable framework** for extracellular matrix quantification, with implications for biomarker discovery, computational pathology, and integration of stromal features into digital health platforms.

## Introduction

Second Harmonic Generation (SHG) signals arising from the highly polarizable, ordered, non-centrosymmetric fibrillar structure of collagen were first reported nearly four decades ago [1]. Since then, SHG imaging has emerged as a powerful optical approach for investigating collagen organisation and deposition in biological tissues [2,3]. Because SHG selectively detects fibrillar collagen without the need for exogenous labelling, it enables clear distinction between fibrillar collagen and surrounding non-fibrillar tissue components. This capability has enabled quantitative assessment of collagen deposition and architecture in several disease contexts. For example, SHG has been used to quantify fibrillar collagen deposition in liver fibrosis for prognostic evaluation [4,5].

In cancer biology, SHG imaging has revealed clinically relevant features of the extracellular matrix. In breast cancer, SHG-derived Tumour Associated Collagen Signatures (TACS) have been identified as independent prognostic indicators of disease-free survival [6,7]. Similarly, in pancreatic ductal adenocarcinoma, SHG-derived collagen deposition scores have been shown to correlate with overall patient survival [8]. These studies highlight the importance of collagen organisation and deposition as clinically informative features of the tumour microenvironment.

Despite these advantages, the application of SHG microscopy has largely remained confined to research settings. The high cost of instrumentation, requirement for specialized expertise, relatively limited field of view, and the time required for tissue imaging pose practical barriers to its implementation in routine diagnostic workflows [9]. In contrast, collagen-specific histochemical stains such as Masson’s Trichrome, Picrosirius Red, and immunohistochemical detection using collagen-specific antibodies are routinely used in clinical pathology to visualize and report collagen deposition and stromal architecture. With the increasing adoption of digital pathology, these stained tissue sections can now be digitized at high resolution and computationally analysed to extract quantitative features of collagen organisation.

Recent studies have therefore begun exploring whether collagen features quantified from digitized histopathological images of stained tissue sections can compare to the information obtained from SHG microscopy, thereby offering a more accessible route for collagen analysis in clinical settings. However, systematic comparisons between SHG-derived collagen measurements and those obtained from routinely stained histopathological sections remain limited.

In this study, we investigate whether collagen features quantified from digital images of Masson-Goldner’s Trichrome-histopathological sections are comparable to those derived from SHG imaging. Because SHG-based collagen quantification is conventionally performed on unstained tissue sections, with only limited reports on stained sections, we first evaluated the compatibility of SHG microscopy for collagen fibre detection in trichrome-stained slides to determine whether trichrome staining interferes with SHG-based collagen detection. The SHG signals obtained from trichrome-stained sections were compared with those from corresponding unstained and hematoxylin and eosin (H&E) stained tissue sections.

Following validation of SHG compatibility with trichrome-stained sections, the same slides were digitized using whole-slide imaging. Matched regions of interest (ROIs) corresponding to the SHG-imaged areas were identified and subjected to quantitative collagen analysis using two independent image analysis approaches: a conventional ImageJ-based workflow and a machine learning (ML)-based computational pipeline.

In summary, our study demonstrates that collagen structural features quantified from digitally scanned Masson-Goldner’s Trichrome-stained histopathological sections show strong concordance with measurements obtained from SHG microscopy. We further show that SHG imaging is compatible with trichrome-stained tissue sections, enabling direct comparison between the two imaging modalities on the same tissue regions. Computational analysis of whole-slide digital images using both conventional image analysis and machine learning-based pipelines was able to capture detailed collagen fibre features and spatial patterns comparable to those derived from SHG imaging. Importantly, the digital pathology-based approach offers practical advantages including larger field-of-view, compatibility with routine clinical staining protocols, and greater scalability for high-throughput analysis. Together, these findings suggest that quantitative analysis of collagen features from routinely acquired histopathological images may provide a feasible and clinically accessible alternative for assessing collagen organisation in tumour tissues.

Further, we systematically evaluated whether collagen structural features quantified from digitally scanned histopathological sections stained with Masson-Goldner’s Trichrome can recapitulate measurements obtained from SHG microscopy. We first assessed the compatibility of SHG imaging with trichrome-stained sections by comparing collagen fibre detection across unstained, H&E-stained, and trichrome-stained tissue sections. Following this validation, matched regions of interest from the same tissue sections were analysed using whole-slide digital imaging and two independent computational pipelines: an ImageJ-based workflow and a machine learning-based analytical framework. Through this comparative analysis, we examine the extent to which collagen features derived from routine histopathological imaging can approximate those obtained through SHG microscopy and evaluate the potential of digital pathology-based approaches as scalable and clinically implementable alternatives for quantitative collagen analysis in tumour tissues.

## Methods

### Patient tissue samples

Primary breast tumour tissue (Formalin-fixed paraffin embedded, FFPE) was received from the biobank [10], with appropriate patient consent and ethical approval. Ethical approval was given for the project titled “Molecular profiling of Indian Triple-Negative Breast Cancer”, on September 30^th^, 2022, by a registered Independent Ethics Committee (Registration EC/NEW/INST/2021/2443 Dated 12/May/2022). Two IDC and one benign tumour sample were selected for the study purpose.

### Sectioning of FFPE Tumour Blocks

FFPE blocks of the selected samples were processed for histopathology. Tumour sections of 3–5 μm were obtained using Leica Microtome RM2255.

### Processing of Unstained Sections

Deparaffinised and rehydrated tumour tissue sections were gradually dehydrated in ethanol solutions followed by Xylene. Slides were then mounted in DPX (catalogue number: Q18404).

### H&E staining

Tissue slides were deparaffinized and gradually rehydrated with ethanol solutions further each slide was cleaned with distilled water and stained with a drop of undiluted Hematoxylin solution (Delafield, catalogue number: 38,803) in a humidifying chamber for 15 min. Slides were then rinsed and subjected to an acid-alcohol wash and immediately transferred to a running water bath 15 mins. This was followed by 1% eosin (Qualigen, catalogue number: Q39312) staining for 30 secs. The slides were then gradually dehydrated in ethanol solutions followed by Xylene. Slides were mounted in DPX (catalogue number: Q18404).

### Trichrome Staining

For Masson Goldner’s Trichrome staining, deparaffinised and rehydrated slides were incubated in Bouin’s solution at 56°C for one hour. Following a wash, they were stained with prepared Weigert’s iron hematoxylin (Hematoxylin - Sigma-Aldrich, catalogue number: H3136-25G) for ten minutes at room temperature and then kept in running water for 5 mins. Slides were then sequentially stained with a drop of Azophloxine (Millipore, catalogue number: 273742) for 7 mins, Tungstophosphoric Acid Orange G solution (Millipore, catalogue number: 273744) for 6 mins, and Light Green SF (Millipore, catalogue number: 273745) for 8 mins. These were interspersed with a 30 sec 1% acetic acid wash. The slides were then gradually dehydrated in ethanol solutions followed by Xylene. Slides were mounted in DPX (catalogue number: Q18404).

### WSI acquisition of stained slides

Whole Slide Imaging (WSI) was done for each slide at 400X by OptraScan using OS-15 bright field digital scanner. Images were checked for focus and quality, converted to TIFF format. WSI TIFF files were uploaded to PathPresenter®. Regions of interest for SHG imaging were annotated.

### Experimental setup for SHG and TPEF Image acquisition of stained and unstained slides

An optical parametric oscillator (APE Levante-IR) was pumped by a fixed wavelength femtosecond pulsed laser source (Fidelity-HP) centred at 1040 nm to generate a signal beam in the wavelength at 1600 nm with 200 fs pulse width and 80 MHz repetition rate. The signal beam at 1600 nm was subsequently directed to a harmonic generator (APE HarmoniXX SHG) to generate a harmonic wavelength at 800 nm, which was used as the source of excitation to acquire second harmonic generation (SHG) and two photon emission fluorescence (TPEF). The sample was mounted in an inverted position on a x-y stage of the inverted optical microscope (Olympus IX73). A water immersion objective (60x/1.2 NA) was used to focus the input laser onto the sample to collect the backward emitted SHG signal at 400 nm. The SHG and TPEF images from the breast tissue samples for same field of view was collected separately using a photomultiplier tube (Hamamatsu R3896) with two different filter sets of wavelength range 400 ± 40 nm and 520± 40 nm, respectively. The incident power at the focus for the tissue samples was 1.75 mW. The sample images are acquired by scanning the laser beam with a pair of galvanometric mirrors (Thorlabs GVS002). In the first round of imaging, three 100 × 100 µm^2^ FOVs were captured referencing the ROI annotations for the IDC High Collagen sample. In the second round of imaging, 25 images each of size 250 × 250 pixels of 0.44 µm pixel size with an overlap region of 10 µm were acquired and stitched to generate a ROI of size 500 × 500 µm^2^. The diffraction limited optical resolution is ∼300nm. Two such ROIs were imaged for each tissue sample, namely benign tissue, IDC tissue with low collagen deposition and IDC tissue with high collagen deposition.

### SHG Image Analysis

The SHG images from Trichrome stained slides were analysed using imageJ to measure collagen deposition patterns. Analysis was done for twenty-five FOVs of 110 µm x 110 µm dimension each sample as well as the stitched ROIs of 500 µm x 500 µm dimension. The acquired SHG images were processed in MATLAB and ImageJ to extract key quantitative parameters including mean SHG signal, and fibre width. Built-in MATLAB image processing filters were applied to suppress noise, and histogram equalization was used to enhance the contrast of the raw images. Frangi filter was applied onto the contrast enhanced image to sharpen the edges of the fibres. Binarization was done on the contrast enhanced image with and without frangi filter using Otsu’s method [11]. The resulting binary images were combined using logical OR operation to retrieve all the fibril pixels represents the complete fibril map. The combined mask was subsequently multiplied with the raw image to isolate fibril-associated intensities, enabling quantification of the mean SHG signal. This signal, indicative of the structural organisation and ordering of collagen within the field of view, was computed as the average SHG intensity over pixels corresponding to the segmented fibrils. In parallel, the binary mask was applied to the contrast-enhanced image to extract fibril structures for morphological analysis. The extracted fibres from the contrast-enhanced images were further analysed using a ridge detection algorithm in ImageJ to estimate the fibre width.

For the calculation of collagen area, SHG images underwent denoising, contrast enhancement and binarization. The percentage of collagen area was measured from these binary images. The pre-processed images were also used as inputs for the TWOMBLI macro on ImageJ to get endpoints, branchpoints and coherence measurements [12]. Orientation frequencies of collagen fibres from 500 µm ROIs were measured using ImageJ’s OrientationJ plugin [13] with a local window of 2 pixels, cubic spline gradient and minimum energy 10%.

### Digital Image Analysis with TWOMBLI

ROIs corresponding to the SHG images were extracted from the digital brightfield image of the trichrome stained WSI using ImageScope software by aligning identifiable objects in the image (S1 Fig). These ROIs were 1716 ×1716 pixels^2^ with a resolution of 0.233 µm, thus with a height and width of 400 µm that corresponded to 500 µm dimensions of an SHG ROI. The digital image ROIs were then deconvoluted in QuPath [14] and further binarized using auto thresholding in ImageJ, area was measured. Binarized images were skeletonised using the TWOMBLI macro on ImageJ. Quantification of endpoints, branchpoints, coherence, and box count was done using this macro. Separately, the 400 µm ROIs were further cropped into 25 FOVs of 80 µm in height and width for each ROI, to correspond to individual FOV images on SHG. These 25 FOVs per image were also analysed using TWOMBLI analysis pipeline. For getting one to one concordance of FOVs between SHG images and digital images, the direction of snake sorting numbering was flipped on the digital image, as digital ROIs were mirror images of SHG ROIs. Number of fibres were calculated as described in Wershof *et al.*, 2021 [12]. The spatial parameters of collagen fibres analysed from the 80 µm patches were plotted for each tissue sample. Orientation frequencies of collagen fibres from 500 µm ROIs were measured using ImageJ’s OrientationJ plugin (citation) with a local window of 10 pixels, cubic spline gradient and minimum energy 10%.

### Digital Image Analysis with ML

ML analysis employs a sophisticated python-based multi-step approach to isolate collagen. It begins with Gaussian smoothing to reduce noise, followed by Mini-Batch K-Means clustering to group pixels based on their RGB colour profiles. A dynamic background detection logic then identifies collagen clusters by analysing brightness and colour neutrality, ensuring that even faint fibres are accurately captured while minimizing false positives from non-fibrous areas.

To detect collagen, corrected stain images were processed using a convolutional neural network (CNN)-based U-Net model. This architecture provided a robust framework for segmenting fine collagen fibres by leveraging high-resolution patch-level inputs. Preprocessing techniques, including gamma correction and Gaussian filtering, were applied to enhance image quality and support reliable feature extraction.

Following collagen detection and segmentation, the extracted fibres were analysed to compute several key metrics characterizing their structural and morphological properties. Circularity statistics were calculated to evaluate the angular orientation of fibres, including the mean circular angle (cir_mean) and its variance (cir_var), which represented the directional consistency of the fibres. Area statistics comprised number of pixels positive for collagen, alongside the total number of fibres (num_fib) detected. Together, these metrics provided a comprehensive statistical framework for characterizing collagen architecture and its spatial organisation.

### Comparison of collagen parameters quantified from two different modalities with three analysis pipelines

Spatial parameter outputs for collagen deposition area, fibre count, Fibre orientation were calculated from both imaging modalities for 25 FOVs from 2 ROIs from each tumour sample type. The outputs from SHG image analysis, brightfield image analysis with Image J pipeline and brightfield image analysis with ML analysis pipeline were compared with each other for concordance using Spearman Correlation analysis. Histograms of orientation frequency distribution from SHG and digital images for each ROI were plotted together comparison.

### Statistical Analysis

All statical test were performed using GraphPad Prism version 8.0.1 for Windows with a significance threshold (alpha) at 0.05. Non-parametric tests were performed since data was not normally distributed. For comparison of means of quantified collagen parameters across three types of samples Kruskal-Wallis test was performed along with Dunn’s multiple comparison test for multiple comparisons. Multiplicity corrected p values were reported for Dunn’s multiple comparison test. Correlation between quantified parameters from different image types and analysis methods was checked using two-tailed Spearman Correlation.

## Results

### SHG Signal detection from three distinctly processed tissue sections

One benign tumour sample and two IDC tumour samples, one with low collagen and another with high collagen deposition were selected for the study. Three sections were of 3 to 5 µm were obtained from each sample. One section of each sample was kept unstained, and the others were stained with H&E and Trichrome staining (Fig 1A). To verify the presence of collagen fibres and to check for variations in signal due to different sample processing, three different ROIs of 100 µm were imaged for SHG and TPEF from unstained, H&E and Trichrome stained slides each for an IDC tumour with high collagen deposition. The sample was imaged with the SHG and TPEF channels through same non-linear optical microscope setup (Fig 1A). A strong SHG signal was observed from the unstained slide indicating presence of collagen fibres with high degree of alignment (Fig 1B). A TPEF signal was also observed from both extracellular and cellular features, however the magnitude of this signal was lesser than that of the SHG signal (Fig 1B). Combined image of SHG and TPEF highlights the fibrillar collagen. However, there are no optical images for reference from the unstained slide (Fig 1B). For an H&E slide (Fig 1C), SHG and TPEF signal were of comparable magnitude indicating the presence of collagen fibres along with the eosin-stained cellular features, as seen in the optical reference images (Fig 1C). Whereas the SHG signal from the Trichrome stained slide (Fig 1D) showed the highest magnitude compared to that of unstained and H&E slide as well as TPEF signal from the same trichrome stained slide. The signal output from the SHG channel was consistent across the three slides namely: unstained, H&E stained and trichrome stained, with the signal from the Trichrome stained slide being the highest in magnitude. The TPEF signal was an order of magnitude lesser than the SHG signal from all the three slides, except from H&E sample. Hence, SHG signal images, and not TPEF images were used for further analysis of collagen fibre characterisation.

**Fig 1.**
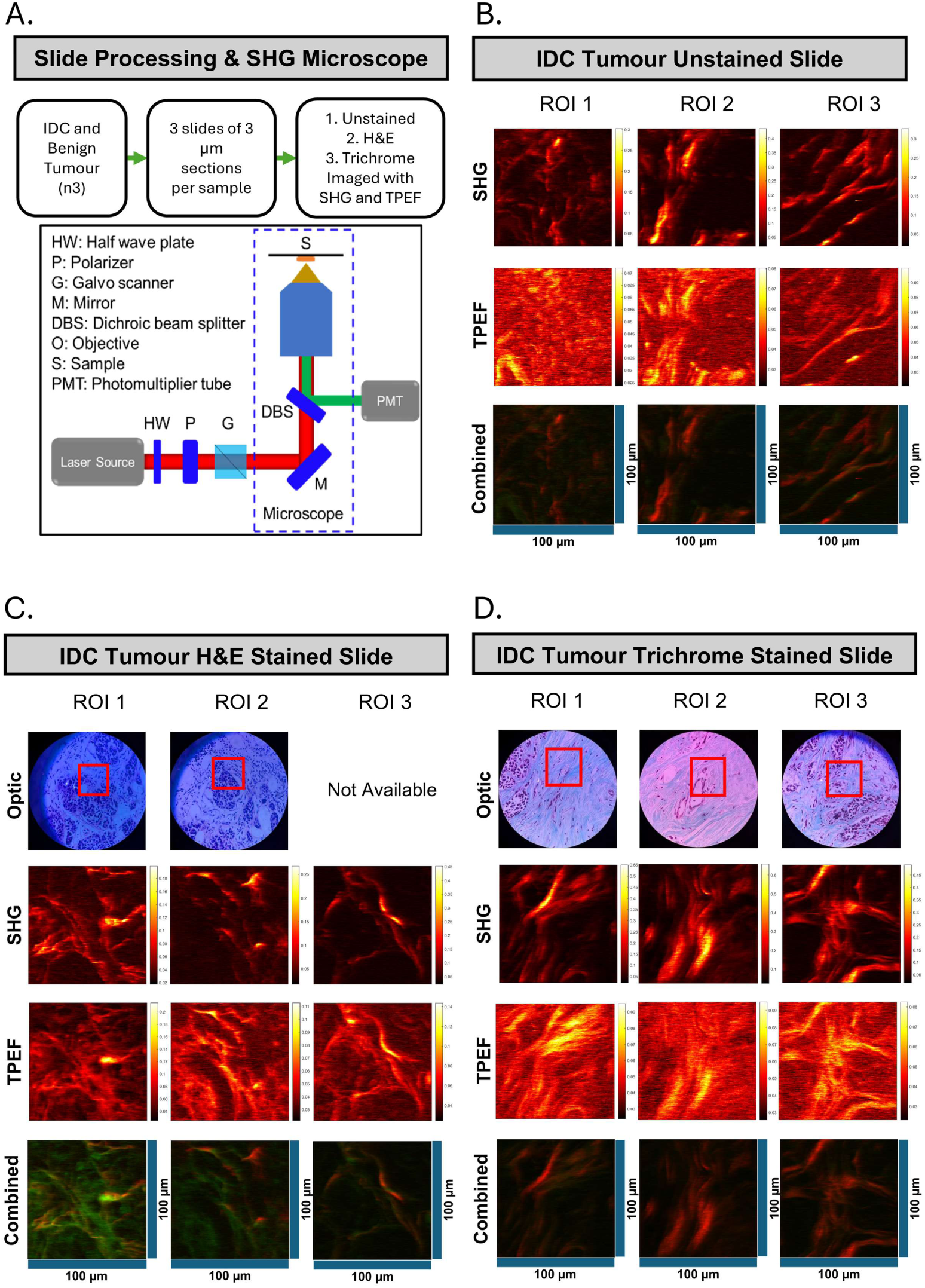
Sample Processing and SHG Signal detection in three tumour samples. (A) Flow chart describing sample processing and imaging with a schematic of the SHG microscope. Three 100 x 100 µm ROIs of the IDC High Collagen sample were imaged using SGH microscopy for (B) Unstained Slides, (C) H&E stained and (D) Masson Trichrome stained Slides. Optical references are shown along with SHG and Two-Photon Excitation Frequency (TPEF) images acquired for the same ROIs for comparison. No optical references are available for the unstained slides. The red signal is from the SHG channel and green from TPEF in the combined images.

### SHG images from H&E and Trichrome Stained tumour tissue samples

Since unstained slides did not provide any visual reference for confirming accuracy of fibre detection, they were omitted from further imaging and analysis. To access larger image region for collagen fibre measurements, twenty-five fields-of-view (FOVs) of dimension 110 µm × 110 µm were imaged and stitched together with a 10 µm overlap to get 500 µm × 500 µm ROIs. Two such ROIs were imaged on H&E stained slides of benign tumour (Fig 2A), IDC tumour with low collagen (Fig 2B) and IDC tumour with high collagen deposition (Fig 2C). ROI to the corresponding SHG image on the left is shown on the bright field image for reference for each SHG image (Fig 2). The SHG signal values for both H&E and Trichrome had comparable magnitudes. We chose to further analyse Trichrome ROIs since Masson’s Trichrome is collagen specific compared to H&E allowing for comparison of spatial metrics of the collagen fibres.

**Fig 2.**
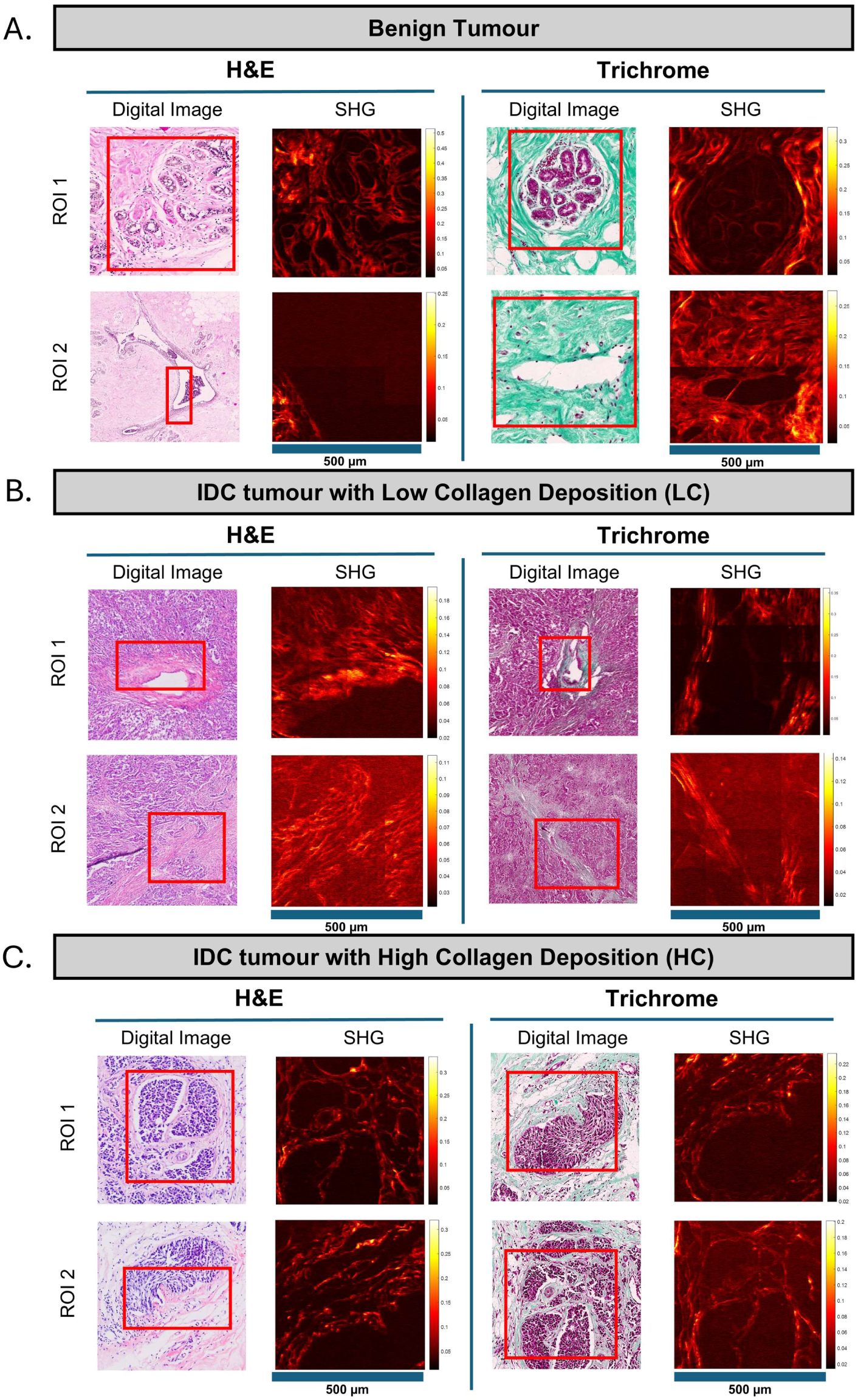
H&E and Trichrome Stained Slides Imaged with SHG Microscopy. Five fields of view (FOVs) with dimensions 110 x 110 µm were imaged with SHG. FOVs were then stitched to generate 500 x 500 µm ROIs. Such two independent ROIs were imaged from H&E and Trichrome Stained Slides each. The two ROIs imaged with SHG, with their corresponding H&E region is shown in (A) for Benign tumour, (B) for IDC tumour with Low Collagen deposition, (C) IDC tumour with High Collagen deposition. Note that all ROIs are imaged at the same on-sample power of 1.75 mW.

### Collagen Fibre visualisation with SHG imaging and Masson Trichrome Stain

Further to compare the two modalities of collagen fibre imaging and quantify collagen parameters, the ROIs imaged with SHG microscopy were identified and cropped out from the digital whole slide image. The 500 × 500 µm^2^ or 1159 ×1159 pixels^2^ stitched ROI of SHG image corresponded to 400 × 400 µm^2^ or 1716 × 1716 pixels^2^ of digital ROI. To digitally identify trichrome stained collagen fibre content, the digital image ROIs were deconvoluted and processed using ImageJ’s TWOMBLI macro. Two ROIs each of digital image, corresponding deconvoluted binary image and the corresponding SHG image are shown for benign tumour (Fig 3A), IDC tissue with low collagen (Fig 3B) and IDC tissue with high collagen (Fig 3C). ROI 1 of the benign tumour sample shows a diffuse deposition of collagen as seen in the digital image. The SHG microscopy image shows loss -of- data with low signal to noise compared with the digital and deconvoluted ROI while SHG image of ROI 2 shows acceptable signal to noise ratio (Fig 3A). For low collagen IDC tissue sample, SHG image showed a considerable amount of noise for ROI2, likely due the low amount of collagen deposition among the cellular structures and was excluded from image analysis (Fig 3B). Both the SHG ROIs for IDC tissue sample with high collagen tissue show high SHG signal output, with less noise when compared to the deconvoluted digital images (Fig 3C). The deconvoluted digital images represented the digital image of the trichrome slides better than the SHG images, with granular details of the collagen fibres intact, indicating minimal data loss.

**Fig 3.**
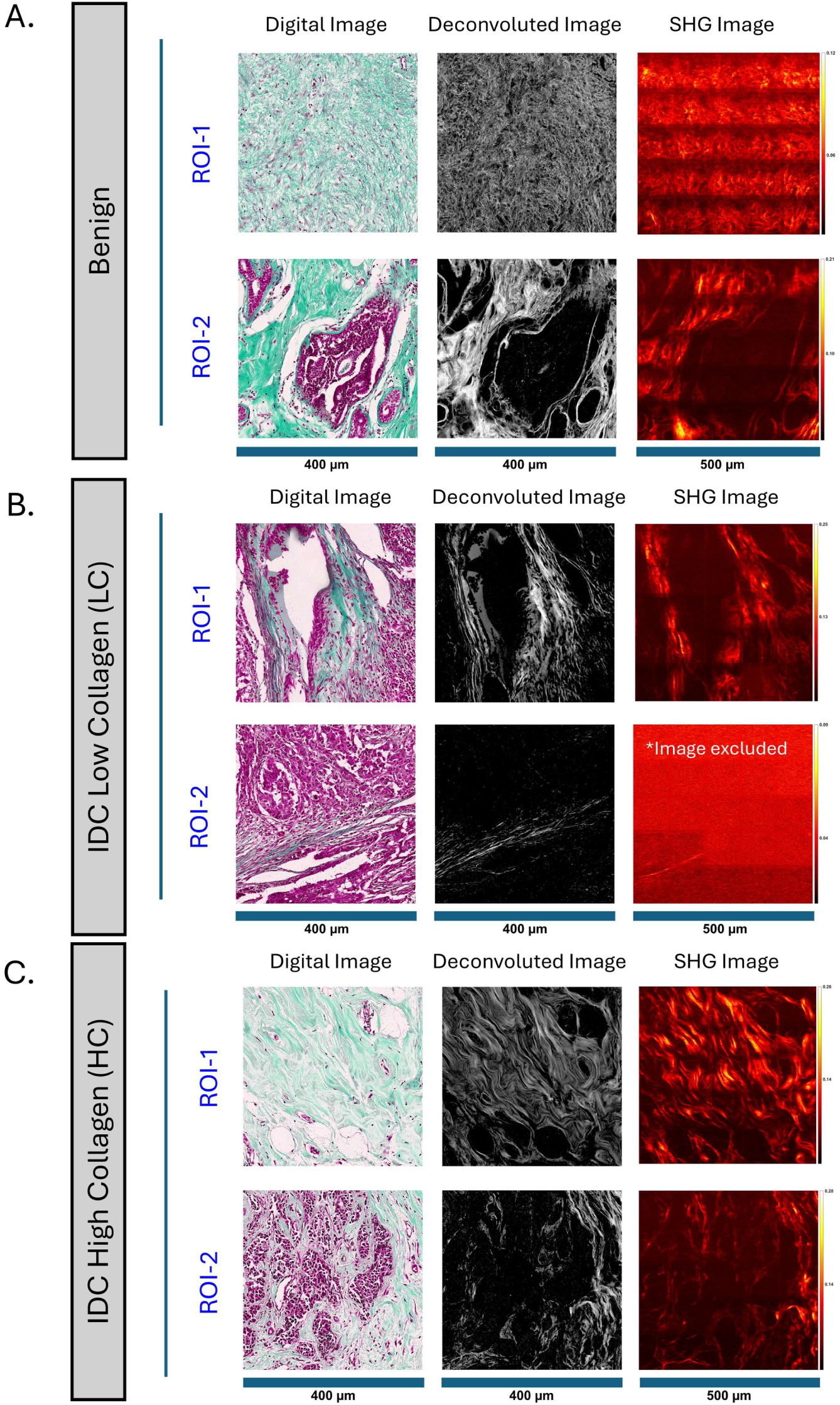
Collagen visualisation with SHG imaging and Masson Trichrome Stain. The same ROIs that were imaged with SHG microscopy were selected from the digital whole slides scanned image of a trichrome stained slide and processed with ImageJ for collagen parameters measurements. Corresponding ROIs of 400 x 400 µm size from the Masson Trichrome images, deconvoluted image of the digital image from the ImageJ pipeline and 500x 500 µm stitched image from SHG microscopy are shown in (A) for benign, (B) for IDC tumour with low collagen deposition, (C) IDC tumour with high collagen deposition. Note that all ROIs are imaged with the same on-sample power. * - IDC tumour with Low Collagen deposition ROI 2 was excluded from analysis due to poor Signal-to-Noise Ratio (SNR).

### Analysis of Collagen Fibre Parameters from SHG microscopy Images

SHG microscopy images were pre-processed and binarized followed by fibril extraction using MATLAB and Image J. SHG signal intensity and fibre width were extracted from the MATLAB and ImageJ scripts, whereas collagen deposition area percentage with respect to total image area, number of fibres, coherency of the fibre alignment were obtained using the TWOMBLI macro. There parameters were quantified from 25 FOVs for each ROI independently (Fig 4A). Mean SHG signal of the FOVs was measured, which is the average SHG signal values of the pixels corresponding to the collagen fibre deposition. The SHG signal differed significantly across the three samples with p < 0.0001 for a Kruskal-Wallis test, with IDC tumour with visual depiction of high collagen indeed showing significantly higher SHG signal corresponding to high collagen deposition compared to benign, p < 0.001, as well as low collagen, p = 0.0002 with Dunn’s Multiple Comparison test (Fig 4B). For fibre width, there was no significant difference observed across samples (Fig 4C). For each tumour slide, percent collagen deposition area did not differ significantly between the two ROIs, across the three tissue samples (Fig 4D). The number of fibres estimated as half the sum of the number of endpoints (E) and branchpoints (B) computed by the TWOMBLI macro [12], differed significantly across the three tissue samples, with a Kruskal-Wallis test p = 0.0235. The number of fibres estimated from two ROIs of IDC tissue with high collagen deposition were significantly higher than those of the IDC tumour tissue with low collagen, p = 0.0224, with Dunn’s Multiple Comparison test (Fig 4E). Alignment of fibres with respect to each other is represented by the coherency metric, yielding values in the range of 0 to 1, with zero representing isotropy while one reflecting perfect alignment [12]. Overall, the coherency differed significantly across the three tissue samples with p < 0.0001 for a Kruskal-Wallis test, while the mean of coherencies of the IDC sample with low collagen was significantly lower than that of the benign sample, p = 0.0002, and high collagen sample, p < 0.0001 with Dunn’s Multiple Comparison test (Fig 4F). The complexity of collagen deposition pattern was represented as box count, ranging from 1 to 2 [12], where one represents a one-dimensional fractal, and two represents more complex fractals filling the space of a two-dimensional plane. The values lesser than 1 represent geometrical dust like object, spaces with finite disconnected points [15]. The quantified box count was lesser than 1 for a considerable number of patches, indicating data loss. Overall, box count differed significantly across the three tissue images, with a Kruskal-Wallis test p = 0.0370, the box count of the benign tumour sample significantly higher than low collagen, p = 0.0324 with Dunn’s Multiple Comparison test (Fig 4G)

**Fig 4.**
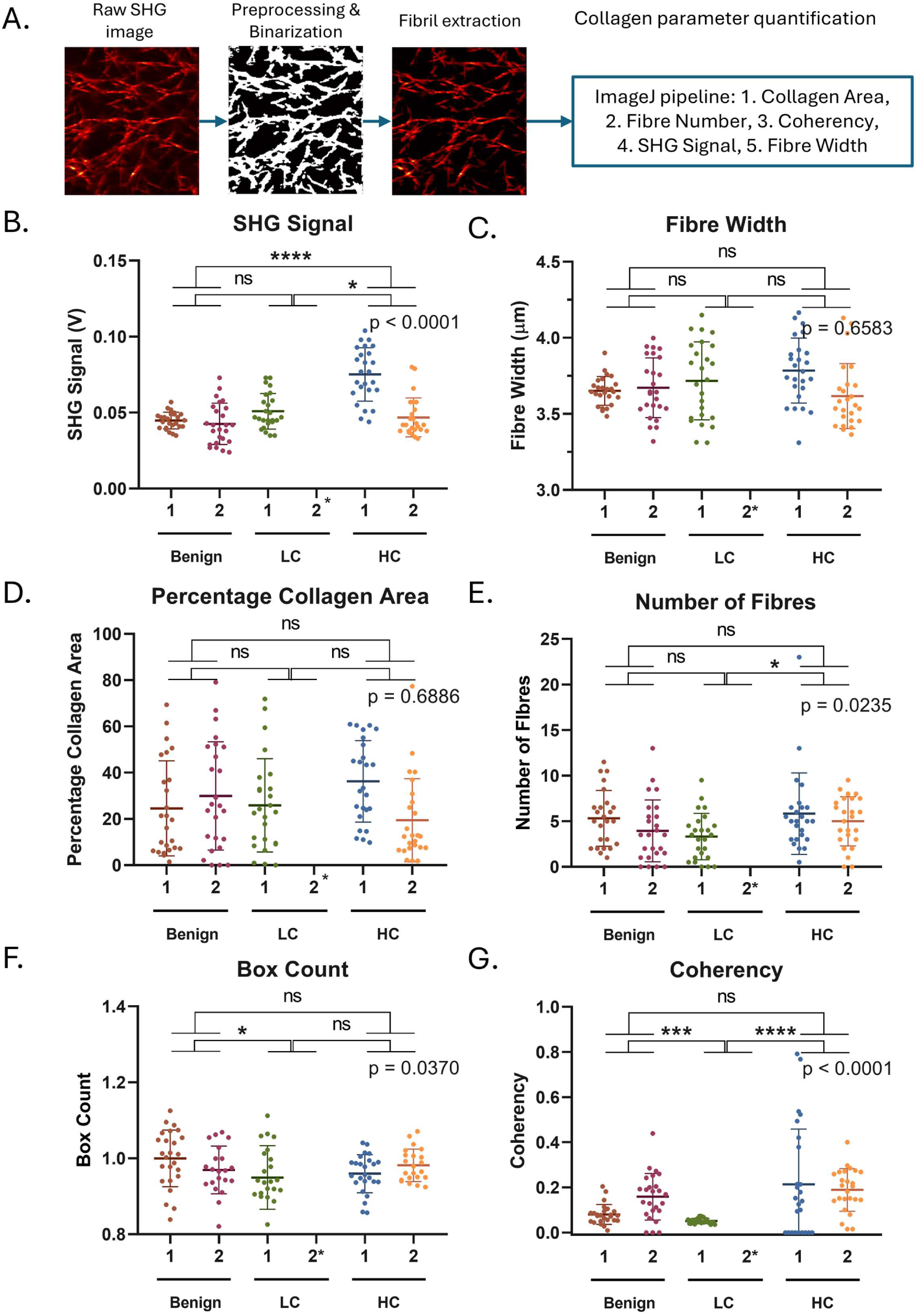
Collagen Architectural Parameters Analysed from SHG Images. (A) Analysis workflow computing of collagen deposition parameters from an SHG Images, Cross-ROI comparison of (B) SHG signal, (C) fibre width. Both SHG signal and fibre width were computed using ImageJ. (D) percent collagen area, (E) number of fibres, (F) box count, (G) coherency were computed from SHG images using TWOMBLI macro. Kruskal-Wallis test performed on the means of both ROIs of each sample. * - ROI 2 of Low Collagen was excluded from analysis.

### Collagen Fibre Parameter outputs analysed from Digital Images

In parallel to SHG microscopy, digital image ROIs were analysed using two independent pipelines (Fig 5A). One pipeline involved analysis using TWOMBLI ImageJ macro [12] where collagen parameters were quantified from deconvoluted binarized FOVs (Fig 5B-E). Another pipeline involved a machine learning (ML) algorithm (Fig 5F-H). From the TWOMBLI pipeline, collagen area percentage differed significantly across the three tissue samples, Kruskal- Wallis test p = 0.0002, with the IDC tumour low collagen sample showing significantly lower collagen area deposition than that of the benign, p = 0.0010, and IDC high collagen tumour sample, p = 0.0011, with Dunn’s Multiple Comparison test (Fig 5B).

**Fig 5.**
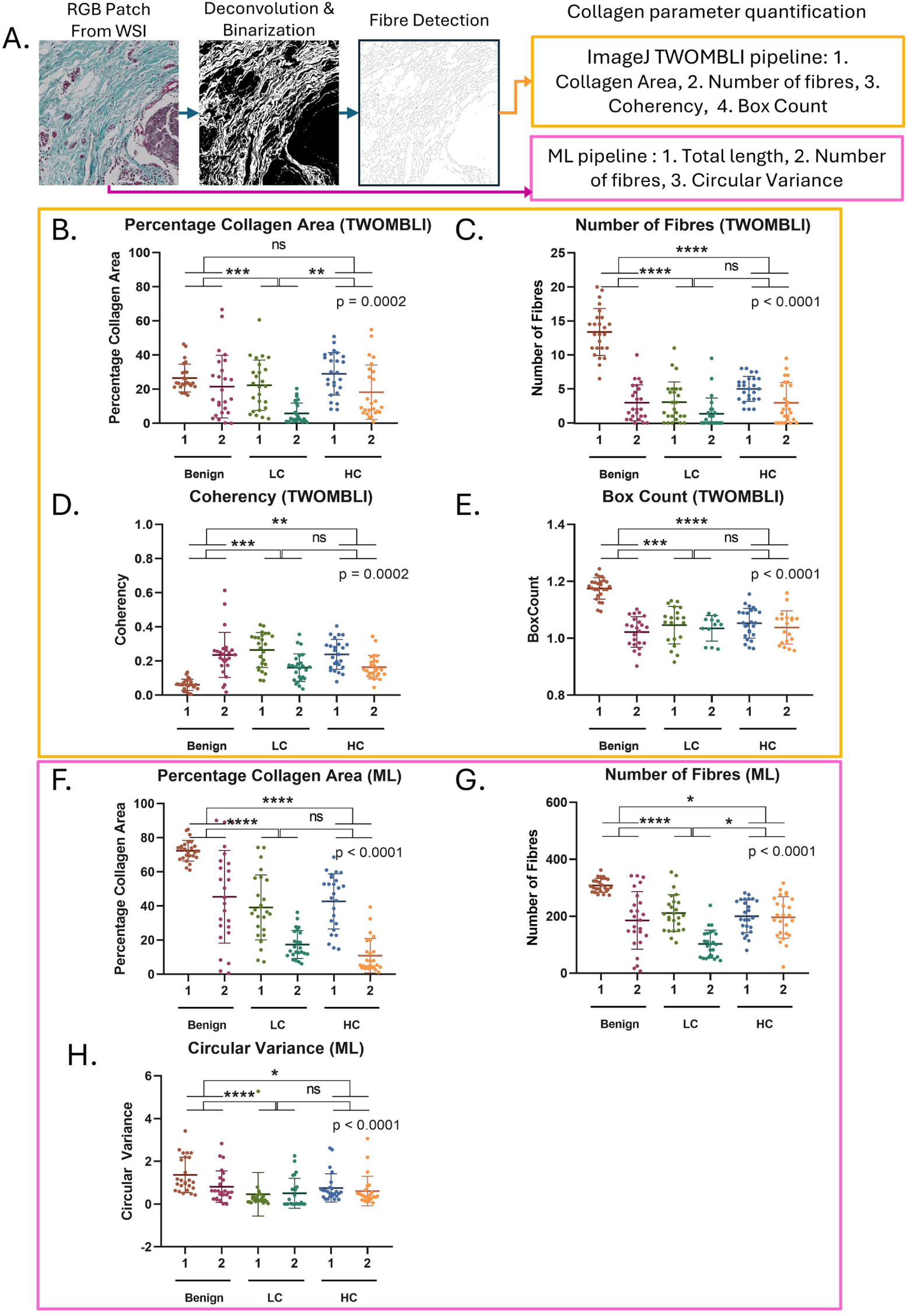
Collagen Architectural Parameters Analysed from Digital Images. (A) Analysis workflow computing of collagen deposition parameters from a digital image, Cross-ROI comparison of (B) percentage collagen area, (C) number of fibres, (D) coherency, (E) box Count quantified using TWOMBLI and (F) percentage collagen area, (G) number of fibres and (H) circular variance quantified using the ML pipeline on the digital image. Kruskal-Wallis test performed on the means of both ROIs of each sample.

The number of fibres were significantly different across samples, Kruskal-Wallis test p < 0.0001, with the benign sample having higher estimate of number of fibres than the low collagen, p < 0.0001, and high collagen sample both, p < 0.0001. Although not significant, high collagen also had higher number of fibres than the low collagen sample, p = 0.0619, with Dunn’s Multiple Comparison test (Fig 5C). Coherency was also significantly different across samples with p = 0.0002 for a Kruskal-Wallis test, p = 0.0002. The benign sample had significantly lesser mean coherency than the low collagen, p = 0.0005, and high collagen sample, p = 0.0018, with Dunn’s Multiple Comparison test (Fig 5D). There was significant difference in the box count of ROIs across samples, p < 0.001 for a Kruskal-Wallis test, with the box count of the benign sample was significantly higher than that of the low collagen, p = 0.0001, and high collagen sample, p < 0.0001, with Dunn’s Multiple Comparison test (Fig 5E).

Overall, the percentage collagen area calculated by ML, indicating amount of deposition, was significantly different across samples, p < 0.0001 for a Kruskal-Wallis test, with more area for the benign sample than that of the high collagen, p < 0.0001 and the low collagen sample, p < 0.0001, with Dunn’s Multiple Comparison test. The percent collagen area for the IDC tumour with high collagen was also significantly higher than the low collagen sample, p = 0.0083 (Fig 5F). The number of fibres from the ML pipeline showed a similar trend to the TWOMBLI estimate significantly differing across samples with a Kruskal-Wallis test p < 0.0001. The benign sample had a higher number of fibres detected than both the high collagen sample, p = 0.0101, and the low collagen sample, p < 0.0001. The number of fibres from the high collagen sample was also significantly higher than the low collagen sample p = 0.0010, with a Dunn’s Multiple Comparison test (Fig 5G). The ML pipeline calculates circular variance. This metric being the variance in the angle of alignment of fibres is a measure of disorder. The trend for circular variance is the inverse of coherency quantified by the TWOMBLI pipeline on the digital images and was significantly different across samples with a Kruskal-Wallis test p < 0.0001. The benign sample had significantly more disorder than the low collagen sample, p < 0.0001, and high collagen sample, p < 0.0001, with Dunn’s Multiple Comparison test (Fig 5H).

### Comparison of collagen deposition quantification with two imaging modalities and three analysis pipelines

Collagen quantification outputs obtained from digital trichrome images using two image-analysis pipelines were compared with measurements derived from Second Harmonic Generation (SHG) microscopy image analysis to assess concordance and reliability. SHG microscopy, which directly detects ordered fibrillar collagen structures, served as the reference standard for collagen quantification. SHG image analysis and the digital trichrome image-analysis pipelines using TWOMBLI generated quantitative measurements of collagen area percentage, fibre number, box count and coherency representing order in alignment of fibres. While the digital trichrome image-analysis pipelines using ML yielded percentage collagen area, number of fibres, and circular variance representing disorder in alignment of fibres. The SHG signal, representing the structural organisation and strength of collagen fibre bundles, was quantifiable only through SHG imaging. Neither of the digital trichrome image-analysis pipelines provided a direct quantitative measure of collagen fibre bundle strength. The closest surrogate metrics available from the digital pipelines were collagen area deposition and fibre number, which reflect collagen abundance and distribution within the tissue and those were compared across the three pipelines along with the alignment metrics as well.

Collagen spatial metrics computed from 25 patches of each of the two ROIs for all three tissue samples were compared across the two imaging modalities and three analysis pipelines using Spearman correlation. The second ROI of the IDC tumour with low collagen sample was excluded from this analysis due to high degree of noise in the SHG image. Between SHG and digital image analysis pipeline using TWOMBLI (Fig 6A-C), there was a significant and strong positive correlation for collagen area percentage with r = 0.6581 and p < 0.001 (Fig 6A), a significant but weak positive correlation for number of fibres (r = 0.2769, p < 0.0001; Fig 6B) and again, a strong positive corelation was observed for weighted coherency as computed by coherency with respect to the collagen area (r = 0.6992, p < 0.0001; Fig 6C).

**Fig 6.**
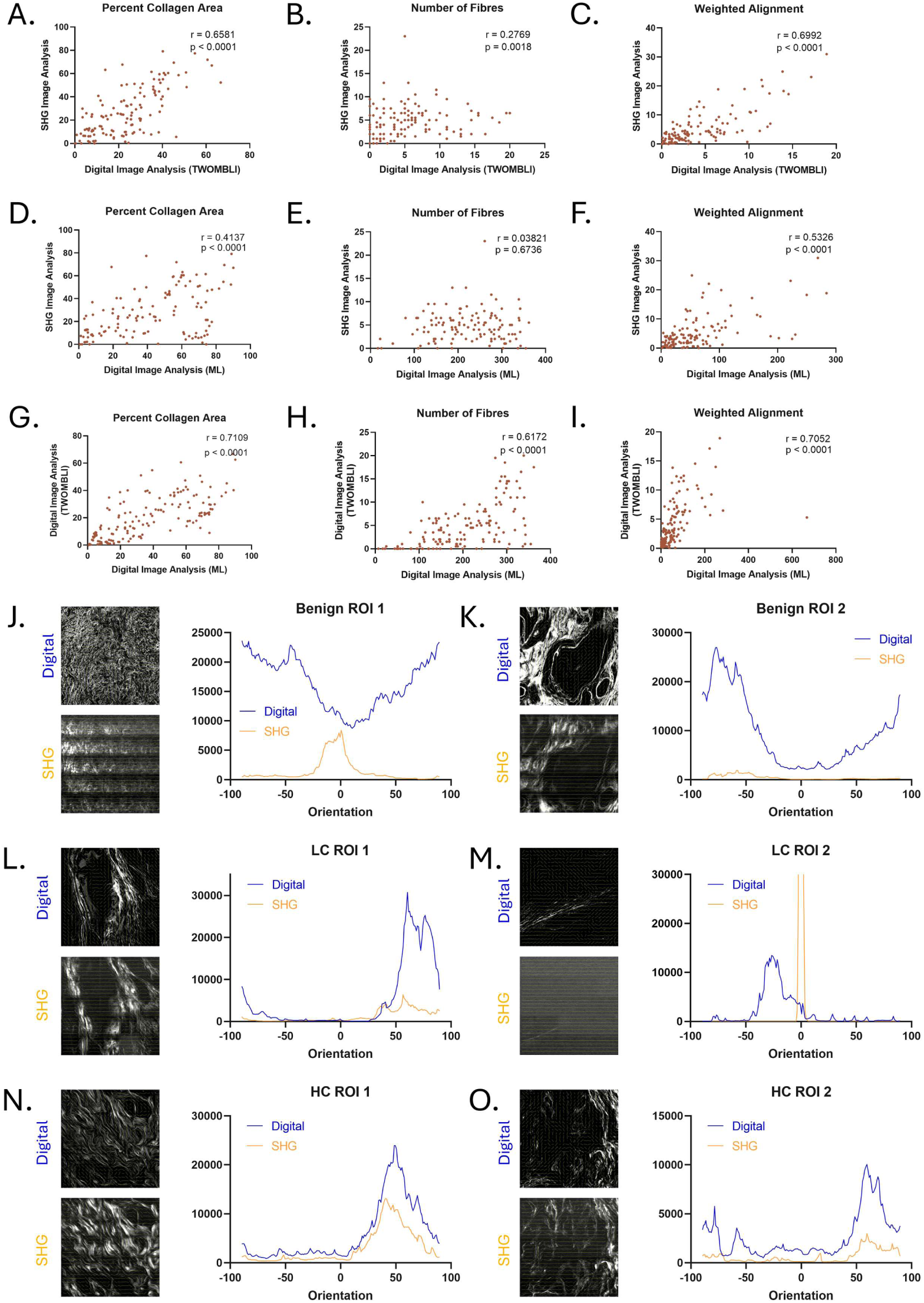
Correlation between Collagen Deposition Parameter measurements from SHG Images and Digital Images. Spearman Correlation of (A) percentage collagen area, (B) number of fibres, (C) weighted coherency measured using TWOMBLI on SHG images and digital Images of Masson Trichrome stained slides. Spearman Correlation of measurements from using TWOMBLI on SHG images and using the ML pipeline on digital images of trichrome stained slides for (D) collagen area % and total length, (E) number of fibres, (F) Weighted Coherency Low Collagen ROI 2 is excluded from the correlation analysis. Spearman Correlation of measurements from digital images of the trichrome stained slide using both the TWOMBLI and ML pipeline for (G) collagen area % and total length, (H) number of fibres, and (I) weighted coherency. Histogram of orientation frequency of collagen deposition with vector fields of the digital and SHG images for (J) Benign ROI 1, (K) Benign ROI 2, (L) Low Collagen ROI 1, (M) Low Collagen ROI 2, (N) High Collagen ROI 1, (O) High Collagen ROI 2. Orientation measurements done on SHG and Digital Images of Masson Trichrome Stained slides.

Following this, metrics quantified using the ML pipeline on digital images and metrics quantified using the TWOMBLI macro on SHG images were compared. Between the SHG image analysis using TWOMBLI and digital image analysis using ML pipelines. there was a significant and positive correlation for collagen area percentage (r = 0.5472, p < 0.0001; Fig 6D) and a weak positive correlation for number of fibres which was statistically significant. (r = 0.1765, p = 0.0489; Fig 6E). Weighted alignment of fibres from the ML metrics is percentage collagen area divided by the circular variance (degree of disorder) and was also positively correlated with the weighted alignment calculated from the SHG image (r = 0.5326, p < 0.0001; Fig 6F). However, the magnitude of these correlations is not as high as the previous comparisons.

Lastly, the digital image analysis pipeline using TWOMBLI and the pipeline using ML was also compared using Spearman correlation. Collagen area percentage showed a significant and strong positive correlation (r = 0.8384, p < 0.001; Fig 6G), number of fibres showed a significant and positive correlation r = 0.6915, p < 0.001; Fig 6H). A significant and positive correlation was seen of weighted alignment (r = 0.7052, p < 0.001; Fig 6G).

Histograms of orientation frequencies were plotted to get a more granular perspective of how orientation measured on digital and SHG images of the trichrome stained slides differs for each ROI. The background vectors for SHG were oriented at 0°, whereas these did not align at a particular angle in the digital images as seen in the vector fields besides the histograms (Fig 6J-O). A concentration of frequencies is distributed around 0° from the SHG image of benign ROI 1, this distribution differs from that of the digital image of the same ROI (Fig 6J). This indicates more noise is being picked up for this ROI, likely due to data loss due to the diffuse nature of collagen deposition. The distribution of the SGH image orientation values slightly aligns with that of the digital image for benign ROI 2, although the magnitude of the digital image orientation is much higher (Fig 6K). The distributions for the low collagen ROI 1 align, with differing magnitudes (Fig 6L). There is a high amount of noise in the SHG image of low collagen ROI 2 as seen by the peak at 0°, whereas the distribution peaks at −25° for the digital image (Fig 6M). The higher noise could be due to the overall low amount of collagen in this ROI. The distributions of orientations from SHG images align strongly with those of the digital images for both ROIs of high collagen (Fig 6N and O).

Overall, the two imaging modalities: SHG microscopy and brightfield digital microscopy images and their analysis pipelines were compared for the data output with its relevance to clinical diagnosis (Table 1). While SHG microscopy has the advantage of directly visualising fibrillar collagen on both unstained and stained tissue sections, the technique necessitates specialised instrumentation and experienced personnel for operation. On the other hand, studying collagen using brightfield microscopy relies on stains that are routinely used by pathologists to visualise clinical fibrosis which involves collagen deposition. The SHG image analysis pipeline demonstrated a higher accuracy of collagen quantification for bundled and aligned collagen fibres, specifically measuring molecular ordering the SHG signal metric. While the brightfield digital image analysis effectively detected broader heterogeneity and was not limited to specific architecture of collagen fibres, quantifying wider range of collagen architectural parameters. Thus, the brightfield images analysed with digital pathology tools may be able to prognosticate a broader range of architecture presenting at the clinic. At a diagnostic setting, a brightfield microscope is routinely available, unlike an SHG microscope. In addition, digital pathology analysis pipeline can be made accessible to path lab computers quite easily in today’s digital age. Ultimately, brightfield analysis of trichrome-stained slides can leverage the scalability and accessibility of established frameworks for clinical investigations.

**Table 1.**
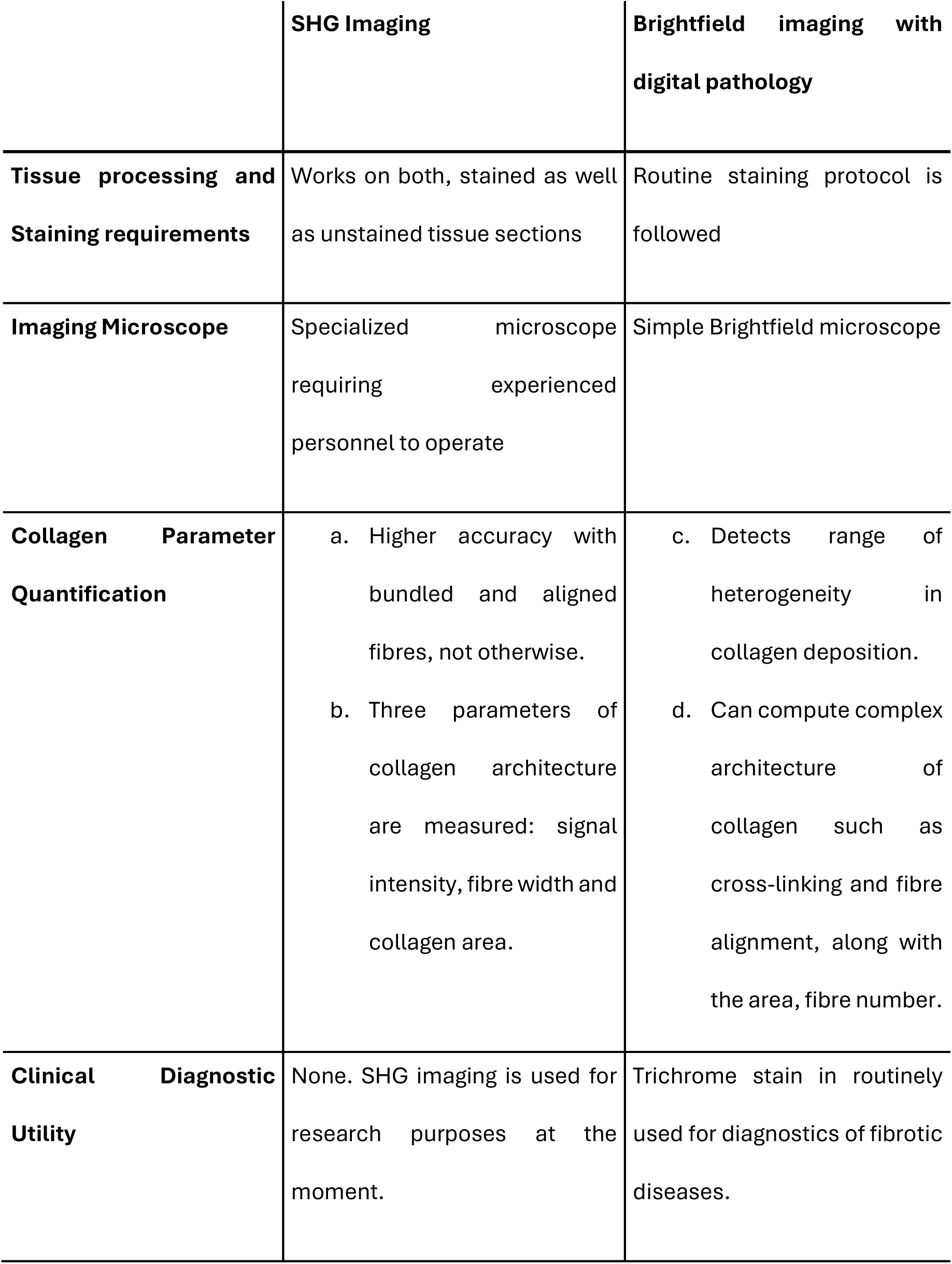
Comparison of two modalities of collagen fibre imaging and measurements.

## Discussion

Collagen organisation within the tumour microenvironment plays an important role in tumour progression, invasion, and patient prognosis. Second Harmonic Generation (SHG) microscopy has emerged as a powerful technique for visualizing and quantifying fibrillar collagen architecture due to its ability to selectively detect non-centrosymmetric collagen fibres without exogenous labelling. However, the high cost, specialized instrumentation, and limited scalability at a diagnostic setting of SHG microscopy restrict its widespread implementation in routine pathology workflows. In this study, we systematically evaluated whether quantitative collagen features derived from digitally scanned Masson-Goldner’s Trichrome-stained histopathological sections can approximate measurements obtained from SHG microscopy, which is considered as a gold standard for collagen architectural measurements.

We first assessed the compatibility of SHG imaging with trichrome-stained sections by comparing collagen fibre detection across unstained, H&E-stained, and trichrome-stained tissue sections. Studies exploring SHG imaging of H&E stained slides have shown that H&E staining is compatible with SHG microscopy [8,16], however it is also reported that the dyes can decrease SHG signal intensity [17] indicating stain interference. A key observation from this study is that SHG imaging remains compatible with trichrome-stained tissue sections. While SHG signals were observed across unstained, H&E-stained, and trichrome-stained sections, and yielded similar collagen features. This observation indicates that trichrome staining does not interfere with collagen SHG detection and in fact may enhance contrast due to improved optical definition of collagen bundles. This compatibility enabled direct comparison of two modalities, namely: SHG imaging with digital brightfield image analysis using the same tissue sections and matched regions of interest with visual reference.

Following this validation, matched regions of interest from the same tissue sections were analysed using whole-slide digital imaging and two independent computational pipelines, an ImageJ-based workflow and a machine learning-based analytical framework. Through this comparative analysis, we examined the extent to which collagen features derived from routine histopathological imaging can approximate those obtained through SHG microscopy and evaluate the potential of digital pathology-based approaches as scalable and clinically implementable alternatives for quantitative collagen analysis in tumour tissues.

Through careful alignment of tissue features, we extracted ROIs from the WSIs of the trichrome stained slides corresponding to the ROIs imaged using SHG from the same slide, making an ROI-to-ROI comparison possible. We observed that although the SHG signal output was satisfactory in the presence of bundled collagen fibres, the deconvoluted digital image of all samples was a better representative of the collagen fibres seen in the digital image than the SHG image. This may be because pixel size of SHG imaging (0.44 µm per pixel) is more than the digital image and the deconvoluted image (0.233 µm per pixel) along with the reduced noise from deconvolution of the digital image. It is worthy to note the high resolution of the digital scanner obtained a matter of minutes, indicating an improvement in brightfield microscopy technology of histological tissue.

Quantitative analysis demonstrated that collagen structural parameters derived from digital trichrome images show strong concordance with SHG-derived measurements. Metrics describing collagen abundance and spatial organisation, including collagen deposition area, fibre number, and fibre alignment, showed statistically significant correlations across imaging modalities. In particular, collagen area and fibre alignment exhibited strong positive correlations between SHG analysis and digital image analysis using the TWOMBLI pipeline [12]. These findings indicate that digital histopathological images can capture spatial characteristics of collagen architecture that are comparable to those obtained using SHG microscopy. Furthermore, the quantifications from the digital image captured the differential presentation of collagen deposition across ROIs. while SHG images showed loss of data loss at lower ranges of collagen deposition.

Two independent digital image analysis approaches: an ImageJ-based TWOMBLI workflow and a machine learning-based computational pipeline, produced broadly consistent quantitative outputs. Strong correlations between these pipelines for collagen area, fibre number, and alignment-related metrics demonstrate the robustness and reproducibility of collagen quantification from digital histopathology images. The machine learning-based framework further enabled automated segmentation of collagen fibres and quantification of fibre orientation disorder using circular variance, illustrating the potential for scalable and automated collagen analysis in whole slide images.

These findings have important implications in the context of **breast cancer stromal biology**. Increasing evidence indicates that the extracellular matrix, particularly fibrillar collagen organisation, actively regulates tumour behaviour by influencing tumour cell migration, mechanotransduction, and immune cell infiltration. Aligned collagen fibres surrounding tumours have been associated with invasive tumour fronts and poor clinical outcomes [6,7]. Quantitative analysis of collagen architecture therefore represents an important avenue for identifying stromal biomarkers of tumour progression. By demonstrating that spatial collagen metrics extracted from routine histopathological images correlate with SHG-derived measurements, our findings suggest that clinically relevant features of tumour collagen organisation may be captured using widely available digital pathology infrastructure.

A major advantage of the digital pathology-based approach lies in its **greater scalability and compatibility with routine clinical workflows**. The larger field-of-view available through whole slide imaging also enables evaluation of stromal heterogeneity across tumour sections, which is difficult to capture using SHG microscopy alone. As digital pathology platforms become increasingly integrated into diagnostic practice, computational analysis of collagen architecture from routinely stained slides could enable large-scale, standardized evaluation of stromal features across clinical cohorts.

The present study has several limitations. First, the analysis was performed on a limited number of tissue samples, and validation in larger patient cohorts will be necessary to confirm the robustness and generalizability of the observed correlations. Second, differences in spatial resolution between SHG microscopy and brightfield digital images required approximate matching of regions of interest rather than exact pixel-level correspondence. Additionally, while digital image analysis captured several spatial features of collagen organisation, it cannot directly measure nonlinear optical properties of collagen fibres, such as SHG signal intensity, that reflect molecular ordering within collagen bundles.

Despite these limitations, this study provides an important methodological advance by demonstrating that **collagen architectural features derived from routine histopathological images can approximate SHG-derived collagen measurements**. To our knowledge, this work represents one of the few systematic comparisons integrating nonlinear optical imaging, conventional image analysis, and machine learning-based computational pipelines for quantitative collagen analysis within matched tissue regions.

In conclusion, our findings demonstrate that collagen structural features quantified from digitally scanned Masson-Goldner’s Trichrome-stained sections show strong concordance with SHG-derived measurements of collagen organisation. Computational analysis of routine histopathology images using both conventional image processing and machine learning approaches enables scalable quantification of collagen architecture while maintaining consistency with SHG-based measurements. As stromal features increasingly emerge as important biomarkers in tumour biology, digital pathology-based collagen quantification may provide a clinically accessible and scalable strategy for integrating extracellular matrix analysis into routine cancer diagnostics and translational research.

## Data Availability

Quantifications are available in the S1 Dataset as part of Supporting Information.

## Acknowledgements

We would like to formally acknowledge Sampada Ghute and Sakshi Durge for sectioning the FPPE breast tumour samples. We acknowledge Sakshi Durge for carrying out histopathology of the breast tumour sections with H&E.

## Author Contributions

VI and MK conceived the overall research goals and aims with support from VR. VI standardized Masson-Goldner’s trichrome staining, carried out histopathology of FFPE breast tumour sections, validated and applied imageJ-based workflow for digital images. KD standardised, validated and carried out SHG imaging. JKM and KD pre-processed and obtained quantitative metrics from SHG images. SG and NS built and applied the ML model for quantification of digital images. VI carried out formal statistical analysis of data. VR provided the NLO microscope for SHG imaging. NS provided computing resources for ML-based quantification. VI and MK visualised the data and drafted the manuscript for publication. MK, NS and VR supervised the project. MK, CK, and LSS acquired ethical approval for the use of breast tumour samples from the PCCM biobank and financial support for research activities. All authors read and approved the final version of the manuscript.

## Supporting information

**S1 Fig.**
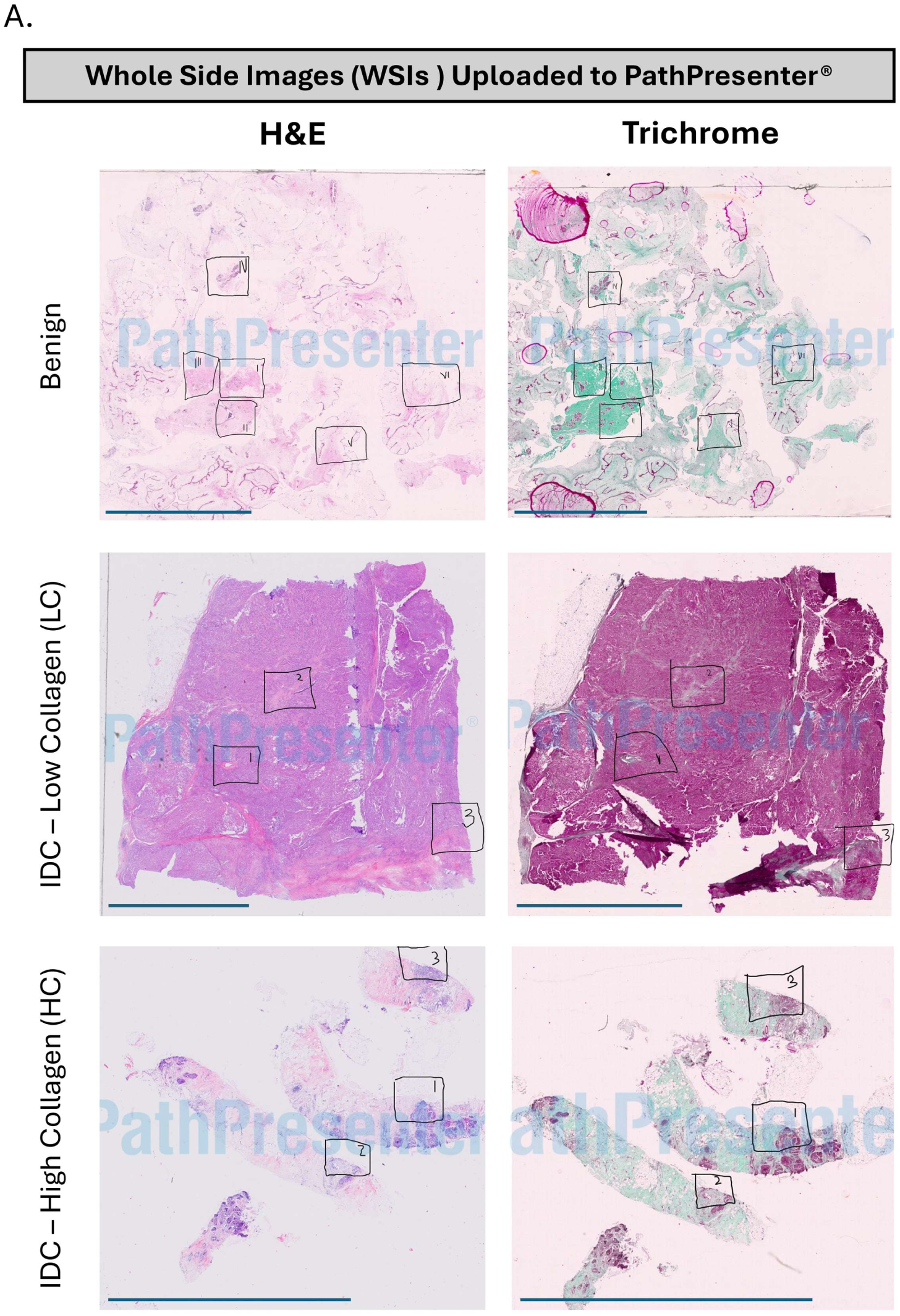
Representative Annotation on Whole Slide Images (WSI). Thumbnail images of whole slide images (WSIs) uploaded to PathPresenter® with annotations marking regions of interest on H&E and Trichrome Stained slides of (A) Benign Tumour, (B) IDC tumour with Low Collagen deposition based on Masson Trichrome stain and (C) IDC tumour with High Collagen deposition, for SHG imaging. Scale bars are 7 mm

**S1 Dataset. Quantifications of collagen fibre parameters.** This file contained measurements of collagen fibre parameters of each patch of the selected SHG and digital ROIs.

